# Dilution-based Evaluation of Airborne Infection Risk - Thorough Expansion of Wells-Riley Model

**DOI:** 10.1101/2020.10.03.20206391

**Authors:** Sheng Zhang, Zhang Lin

## Abstract

Evaluation of airborne infection risk with spatial and temporal resolutions is indispensable for the design of proper interventions fighting infectious respiratory diseases (*e.g*., COVID-19), because the distribution of aerosol contagions is both spatially and temporally non-uniform. However, the well-recognized Wells-Riley model and modified Wells-Riley model (*i.e*., the rebreathed-fraction model) are limited to the well-mixed condition and unable to evaluate airborne infection risk spatially and temporally, which could result in overestimation or underestimation of airborne infection risk. This study proposes a dilution-based evaluation method for airborne infection risk. The method proposed is benchmarked by the Wells-Riley model and modified Wells-Riley model, which indicates that the method proposed is a thorough expansion of the Wells-Riley model for evaluation of airborne infection risk with both spatial and temporal resolutions. Experiments in a mock hospital ward also demonstrate that the method proposed effectively evaluates the airborne infection risk both spatially and temporally.

## 1. Introduction

Infectious respiratory diseases (*e.g*., tuberculosis, influenza, and aspergillosis) are severe threats to people’s health and economic development [1]. Particularly, the global pandemic of COVID-19 results in substantial loss of human lives and jeopardizes human development (social, economic, *etc*.). Airborne infection due to the inhalation of pathogen-laden aerosols is one major transmission pathway of infectious respiratory diseases [2]. Infectors’ coughing, sneezing, talking, and breathing generate tens of thousands of infectious droplets, and most of the generated infectious droplets evaporate into the air as infectious droplet nuclei [3, 4]. A COVID-19 infector can yield infectious droplets with 100,000 virions every minute of speaking [4, 5]. The airborne transmission of the infectious droplet nuclei can occur over a long distance, causing cross infections. For example, Yu et al. [6] found that SARS airborne infections occurred between different rooms and between adjacent buildings. Liu et al. [7] measured the concentration of airborne COVID-19 RNA in isolation wards and ventilated patient rooms, and suggested that COVID-19 could be transmitted via aerosols. More and more evidence reveals the airborne infection risk of COVID-19 [8, 9].

Evaluation of airborne infection risk should take into account the spatially and temporally non-uniform distribution of pathogen-laden aerosols [1]. The spread of pathogen-laden aerosols is significantly affected by the complicated and transient interactions among the respiratory flows and thermal plumes of occupants, and ventilation flow, which results in the spatially and temporally non-uniform distribution of pathogen-laden aerosols [10]. Since occupants generally spend more than 90% time conducting indoor activities, indoor ventilation is one of the most effective engineering solutions for reducing airborne transmission by diluting the pathogen-laden aerosols with pathogen-free air [11, 12]. Airflow patterns of advanced ventilation with non-uniform aerosol distribution can more effectively reduce airborne transmission risk. For example, displacement ventilation and stratum ventilation target diluting the airborne contaminants in the breathing zone rather than the entire room, thus improve the contaminant removal efficiency of the breathing zone by up to 50% [13].

However, it is challenging to evaluate the airborne infection risk with spatial and temporal resolutions in practice. The dose-response model and Wells-Riley model are two methods for quantitative evaluation of the airborne infection risk [14]. Since the dose-response model requires the information which is costly to obtain during experimental and on-site studies, *e.g*., the particle sizes and infectivity (involving medical and microbiological sciences), the dose-response model is less frequently used than the Wells-Riley model [1, 10, 15]. The Wells-Riley model is a simple and quick evaluation method of the airborne infection risk, because it uses the concept of quantum to implicitly consider the infectivity, infectious source strength, biological decay of pathogens, *etc*. [14]. As a result, the Wells-Riley model has been widely used in the studies of infectious respiratory diseases [1, 10, 15]. However, the Wells-Riley model is based on the well-mixed and steady assumption that the distribution of pathogen-laden aerosols is spatially and temporally uniform. With this assumption, the airborne infection risk could be underestimated by the Wells-Riley model, and the interventions for reducing airborne infection risk suggested by the Wells-Riley model could be improper [1, 10, 16]. For example, the Wells-Riley model suggests a larger ventilation rate. However, increasing the ventilate rate might deteriorate indoor air quality due to the potential negative effects of the increased ventilation rate on the contaminant removal efficiency of non-uniform airflow pattern [17].

Rudnick and Milton [18] proposed the concept of rebreathed fraction, and used it to modify the Wells-Riley model for the transient condition. The rebreathed fraction is calculated from the difference between indoor and outdoor CO_2_ concentrations [18]. The rebreathed-fraction model has been well recognized for the airborne infection risk with a temporal resolution [10, 19, 20]. However, the rebreathed-fraction model is also based on the well-mixed assumption, and thus cannot evaluate the airborne infection risk spatially [18]. Numerical simulations (*e.g*., computational fluid dynamics) have been used to provide the spatial quantum concentration of airborne pathogens for the Wells-Riley model to evaluate the airborne infection risk spatially [21, 22]. However, this method is inapplicable to physical experiments because it is impossible to measure the quantum concentration of airborne pathogens for experimental and on-site studies [1, 10, 18]. Although numerical simulations are powerful in epidemical studies, physical experiments are indispensable for reliable results [10]. Thus, there is an urgent need for an evaluation method of airborne infection risk with spatial and temporal resolutions for practical applications.

This study will provide a dilution-based evaluation method of the airborne infection risk for both spatial and temporal resolutions. The proposed model is illustrated in Section 2, and benchmarked by the Wells-Riley model under the well-mixed and steady condition in Section 3, and by the rebreathed-fraction model under the well-mixed and transient condition in Section 4. Experiments of a mock hospital ward served by displacement ventilation are conducted in Section 5 to demonstrate the applicability of the method proposed to evaluate airborne infection risk spatially and temporally.

## 2. Dilution-based airborne infection risk estimation proposed

The concept of dilution is diluting the airborne contaminants with clean air so that the concentration of airborne contaminants is reduced. Dilution is the mechanism of ventilation in reducing airborne infection risk [1]. According to the concept of dilution, the dilution ratio is defined as the ratio between the source concentration to the contaminant concentration at the target position (Equation 1). The dilution ratio can vary among different positions relative to the contaminant source transiently. With the dilution ratio, the quantum concentration of airborne pathogens at the target position is calculated with Equation 2, which is the quantum concentration exhaled by the infector diluted at the target position. The quantum is an infectious dose unit [23], and one quantum is the quantity of pathogens required to cause an infection risk of 63.2% (*i.e*., 1-e^-1^) [19]. For example, the quantum generate rate of a Tuberculosis infector as suggested by Andrews et al. [24] is 1.25 quanta/h, and that of an asymptomatic infector COVID-19 is 142 quanta/h [25]. It is noted that the factor of infectious virus removal in Equation 2 (*φ*) is used to account for the effects of interventions in reducing the airborne infection risk except ventilation [14], *e.g*., facial masks [26, 27] and air purifiers [28] recommended for controlling COVID-19.

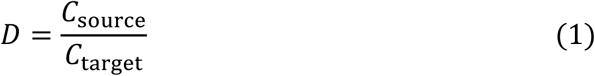

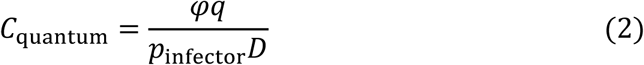

where *C*_source_ and *C*_target_ are the airborne contaminant concentrations at the source and target position respectively (ppm); *C*_quantum_ is the airborne quantum concentration at the target position (quanta/m^3^); *D* is the dilution ratio at the target position; *p*_infector_ is the breathing rate of the infector (m^3^/s); *q* is the quantum generation rate (quanta/s); *φ* is the factor of infectious virus removal.

With the quantum concentration at the target position, the quanta inhaled by a susceptible at the target position over a given exposure period is calculated by Equation 3. With the inhaled quanta, the airborne infection risk of a susceptible at the target position over that exposure period is estimated based on Poisson distribution (Equation 4). The pathogens are discrete matters and distribute in a medium randomly following Poisson distribution [14, 18, 20, 29]. The airborne infection risk is the probability of that susceptible to be infected because of the inhaled airborne pathogens. The dilution-based airborne infection risk proposed is obtained as Equation 5 by combining Equations 2-4.

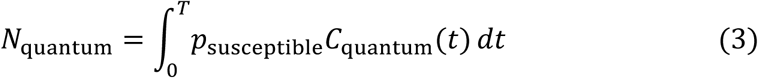

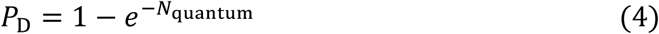

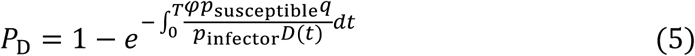

Where *C*_quantum_(*t*) and *D*(t) are the quantum concentration (quanta/m^3^) and dilution ratio at the target position at time *t* during a given exposure period of *T* respectively; *N*_quantum_ is the inhaled quanta by a susceptible at the target position during the given exposure period; *P*_D_ is the airborne infection risk at the target position during the given exposure period estimated by the dilution-based estimation method proposed; *p*_susceptible_ is the breathing rate of the susceptible (m^3^/s), which can be different from that of the infector because the infector and susceptible have different health conditions and might have different activity levels [25].

Since the method proposed employs the concept of the quantum, it has the merit as the Wells-Riley model of implicitly considering the biological complexities of the infectivity, infectious source strength, biological decay of pathogens, *etc*., which makes the evaluation of the airborne infection risk by the method proposed simple and quick [14]. Moreover, compared with the Wells-Riley model, the method proposed has two advantages. First, the method proposed can estimate the airborne infection risk for any target position during any exposure period (Equation 5), *i.e*., the method proposed evaluates the airborne infection risk for both spatial and temporal resolutions. Second, the method proposed is more convenient for practical applications. The Wells-Riley model (Equation 6) requires knowing the numbers of infectors, but the method proposed does not. The information on the numbers of infectors is not always available, particularly when asymptomatic infectors present. It was reported that the asymptomatic infectors were responsible for 79% COVID-19 infections in Wuhan [4, 30]. Besides, the Wells-Riley model (Equation 6) requires the input of the ventilation rate which is inconvenient to measure in practice [18]. For example, Wu et al. [31] measured on-site tracer gas concentration in a residential building, and approximated the ventilation rate from the measured tracer gas concentration for the Wells-Riley model to evaluate the airborne infection risk. The approximation of the ventilation rate from the tracer gas concentration increases the evaluation complexity and decreases the evaluation reliability. In contrast, the dilution ratio can be conveniently and reliably obtained from the tracer gas concentration (Equation 1) for the method proposed to evaluate the airborne infection risk, which will be further demonstrated in Section 5.

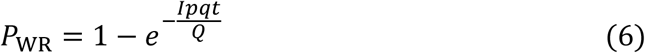

Where *I* is the number of infectors; *P*_WR_ is the airborne infection risk estimated by the Wells-Riley model; *p* is the breathing rate of a typical person (m^3^/s); *Q* is the ventilation rate (m^3^/s); *t* is the time length of the exposure period (s).

## 3. Benchmark with Wells-Riley model under well-mixed and steady condition

As introduced in Section 1, the Wells-Riley model has been well recognized to provide a reliable evaluation of the airborne infection risk under the well-mixed and steady condition. Since the method proposed evaluates the airborne infection risk for both spatial and temporal resolutions, the well-mixed and steady condition is one of the special cases. When the method proposed applies to the well-mixed and steady condition, it should produce identical or similar results to those from the Wells-Riley model. This is proofed as follows. According to the mass conservation law, the variation of contaminants in the space is the generated contaminants in the space minus the contaminants removed from the space by ventilation, which is described by Equation 7 when contaminants are well mixed in the space air [20, 31]. When the contaminants refer to the exhaled air by the infectors (Equation 8) and the contaminants in the ventilation air supply is zero (clean air), the contaminant concentration in the exhaled air is unity and the dilution ratio is the reciprocal of the contaminant concentration in the space air (Equation 9). For the steady condition, the variation of the contaminant concentration in the space air is zero (Equation 10). From Equations 8-10, the dilution ratio under the well-mixed and steady condition is obtained as the function of the ventilation rate, numbers of infectors, and breathing rate of infectors (Equation 11), implying that the dilution ratio implicitly takes into account the ventilation rate, numbers of infectors, and breathing rate of infectors, which makes the implementation of the method proposed convenient in practice as discussed in Section 2. Combing Equations 5 and 11, the method proposed for the well-mixed and steady condition is obtained as Equation 12. In Equation 12, the breathing rate by the susceptible can be the same as that of a typical person and the factor of infectious virus removal can be unity as assumed by the Wells-Riley model. As a result, the proposed model for the well-mixed and steady condition (Equation 12) produces the same airborne infection risk as the Wells-Riley model (Equation 6). This indicates that the method proposed is an expansion of the Wells-Riley model for the evaluation of airborne infection risk with both spatial and temporal resolutions.

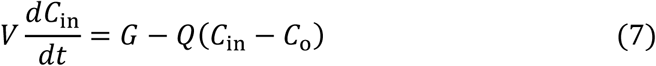

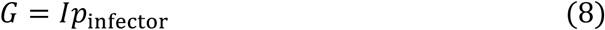

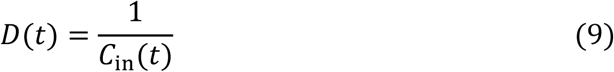

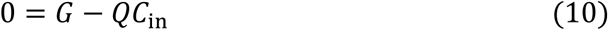

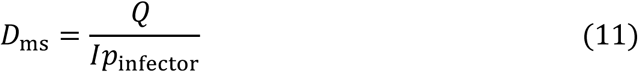

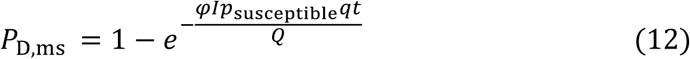

Where *C*_in_ and *C*_o_ are the contaminant concentrations in the space air and ventilation air supply respectively (ppm); *D*_ms_ is the dilution ratio under the well-mixed and steady condition; *G* is the contaminant generation rate in the space (m^3^/s); *P*_D,ms_ is the airborne infection risk estimated by the dilution-based estimation method proposed under the well-mixed and steady condition; *V* is the volume of the space (m^3^).

## 4. Benchmark with rebreathed-fraction model under well-mixed and dynamic condition

The rebreathed-fraction model (Equations 13 and 14) is a modified Wells-Riley model for a reliable evaluation of the airborne infection risk under the well-mixed and transient condition [10, 18-20]. Since the method proposed can evaluate the airborne infection risk for both spatial and temporal resolutions, the well-mixed and transient condition is one of the special cases. When the method proposed applies to the well-mixed and transient condition, it should produce identical or similar results to those from the rebreathed-fraction model. This is proofed as follows. By integrating Equation 7, the transient contaminant concentration in the space air is obtained as Equation 15 [18, 20]. When the contaminants refer to the exhaled air by infectors, the dilution ratio under the well-mixed and transient condition is obtained as Equation 16 from Equations 8, 9 and 15 with the contaminant concentrations in the exhaled air and the ventilation air supply of unity and zero respectively. When the contaminant is CO_2_, the contaminant generation rate in the space is the CO_2_ generated by all occupants (Equation 17), and Equation 15 is transferred to be Equation 18. According to the definition of the rebreathed fraction (Equation 14) [18] and Equation 18, the rebreathed fraction is expressed as Equation 19. From Equations 16 and 19, the relationship between the dilution ratio and rebreathed fraction under the well-mixed and transient condition is obtained as Equation 20. With Equation 20 to replace the dilution ratio in Equation 5, the method proposed under the well-mixed and transient condition is expressed as Equation 21. In Equation 21, the breathing rate by the susceptible can be the same as that of a typical person and the factor of infectious virus removal can be unity as assumed by the rebreathed-fraction model. As a result, the proposed model for the well-mixed and transient condition (Equation 21) produces the same airborne infection risk as the rebreathed-fraction model (Equation 13). This indicates that while the rebreathed-fraction model is a limited expansion of the Wells-Riley model with a temporal resolution of airborne infection risk, the method proposed is a thorough expansion of the Wells-Riley model with both spatial and temporal resolutions of airborne infection risk.

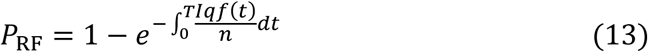

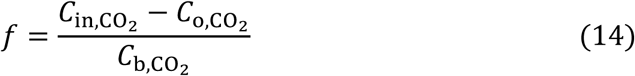

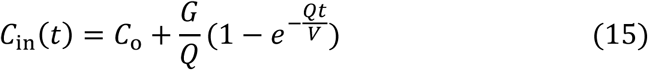

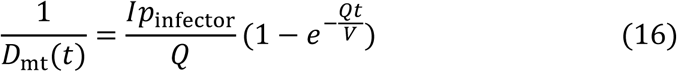

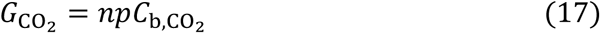

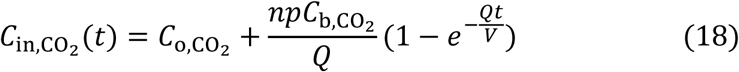

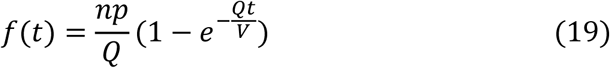

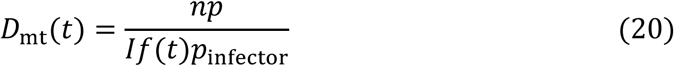

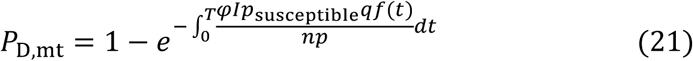

Where 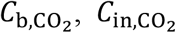, and 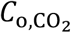 are the CO_2_ concentrations in the exhaled air, space air and ventilation air supply respectively (ppm); *D*_mt_ is the dilution ratio under the well-mixed and transient condition; *f* is the rebreathed faction; *n* is the number of occupants in the space; *P*_RF_ are *P*_D,ms_ are the airborne infection risks under the well-mixed and transient condition estimated by the rebreathed-fraction model and the dilution-based estimation method proposed respectively.

## 5. Demonstration of applicability of method proposed for both spatial and temporal resolutions

Experiments are conducted to demonstrate the effectiveness of the method proposed for the evaluation of airborne infection risk for both spatial and temporal resolutions. The environmental chamber, with dimensions of 8.8 m (length) × 6.1 m (width) × 2.4 m (height), is configured as a mock hospital ward with multiple beds (Figure 1). The ward is served with displacement ventilation that the conditioned air is supplied near the floor level (S1-S14) (with the supply air temperature around 20°C and supply airflow rate around 12 ACH) and exhausted from the ceiling (E1-E3). Displacement ventilation has an airflow pattern with high contaminant removal efficiency at the breathing level [13]. SF_6_ (released from the mouth of the infector in Figure 1) is used as the tracer gas to represent the airborne contaminants [31-33]. Three target positions (L1-L3) with the standing susceptible occupants are concerned, and the tracer gas concentrations at the three target positions are measured at the breathing level (*i.e*., at the height of 1.5 m above the floor). INNOVA 1412i is used to measure the concentration of SF_6_ with a measurement accuracy of 0.06 ppm.

**Fig.1.**
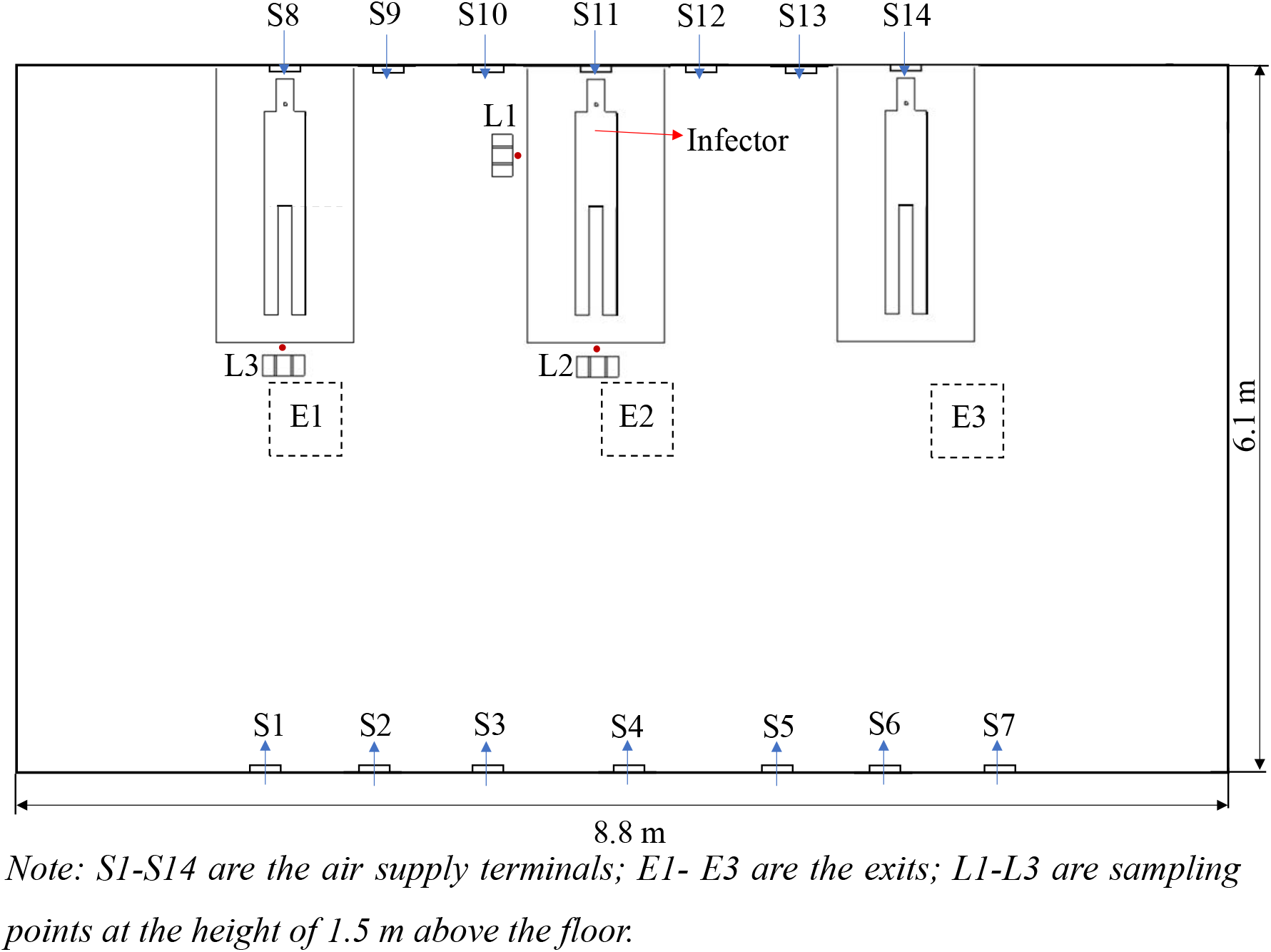
Experimental setup of mock hospital ward with multiple beds served with displacement ventilation.

Figure 2 shows that the tracer gas distribution is both spatially and temporally non-uniform. The concentration of the tracer gas generally increases first and then tends to be steady, but fluctuates slightly due to the randomness induced by air turbulence (Sze and Chao 2010). The tracer gas concentration at the target position of L1 is higher than those at the other two target positions because it is the closest to the infector. From the tracer gas distribution, the dilution ratio is calculated (Equation 1). The dose rate of the tracer gas is 2 ml/s and the breathing rate of the resting infector is 0.49 m^3^/h [25], resulting in a contaminant concentration of the source around 14,700 ppm for the calculation of the dilution ratio (Equation 1). Figure 3 shows that the dilution ratio also has spatial and temporal resolutions, varying from around 2000 to 14,000. The dilution ratios at the three target positions decrease first and then tend to be steady, and the dilution ratio of the target position (L1) closest to the infector is the smallest.

**Fig.2.**
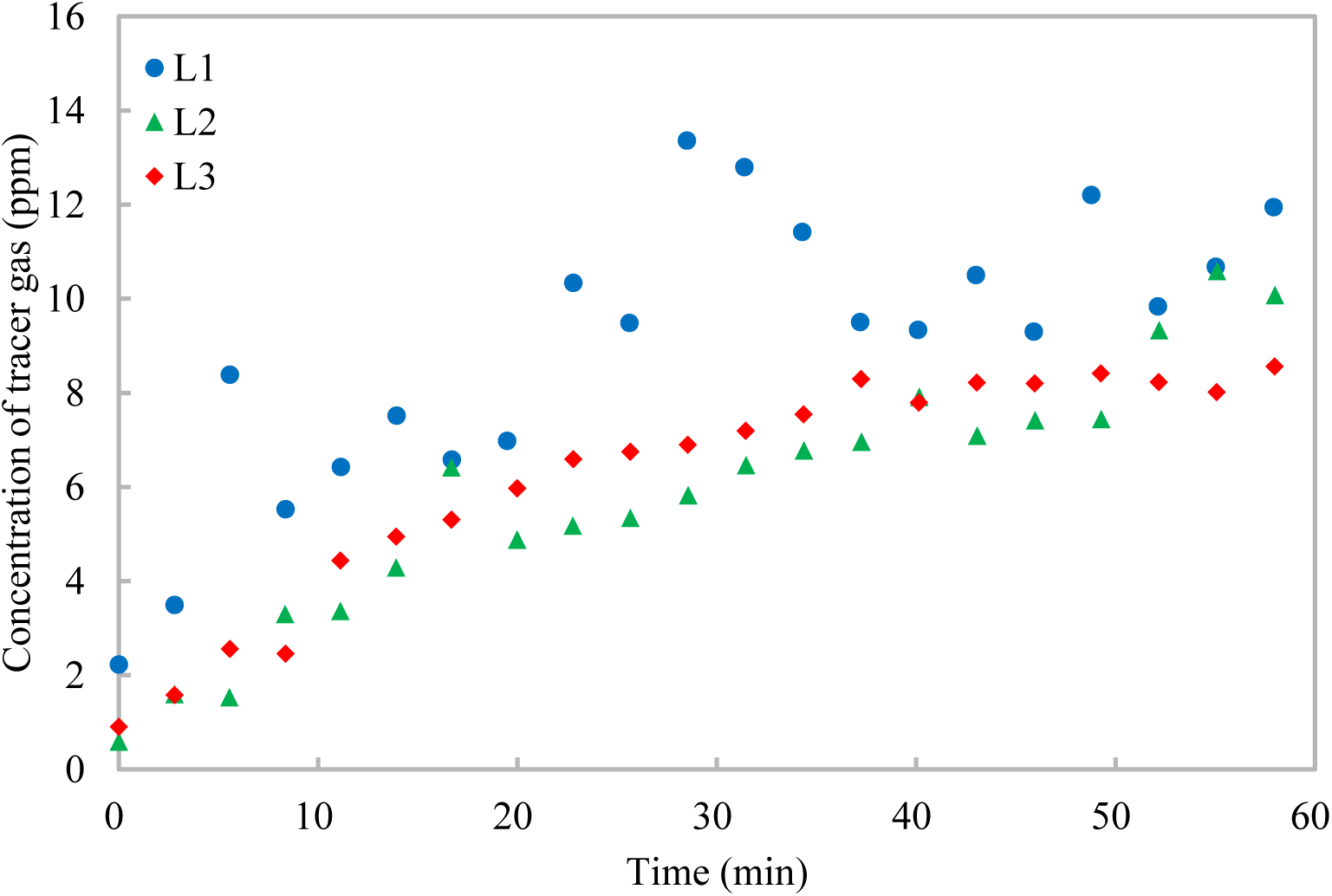
Variations of tracer gas concentrations at different target positions with time.

**Fig.3.**
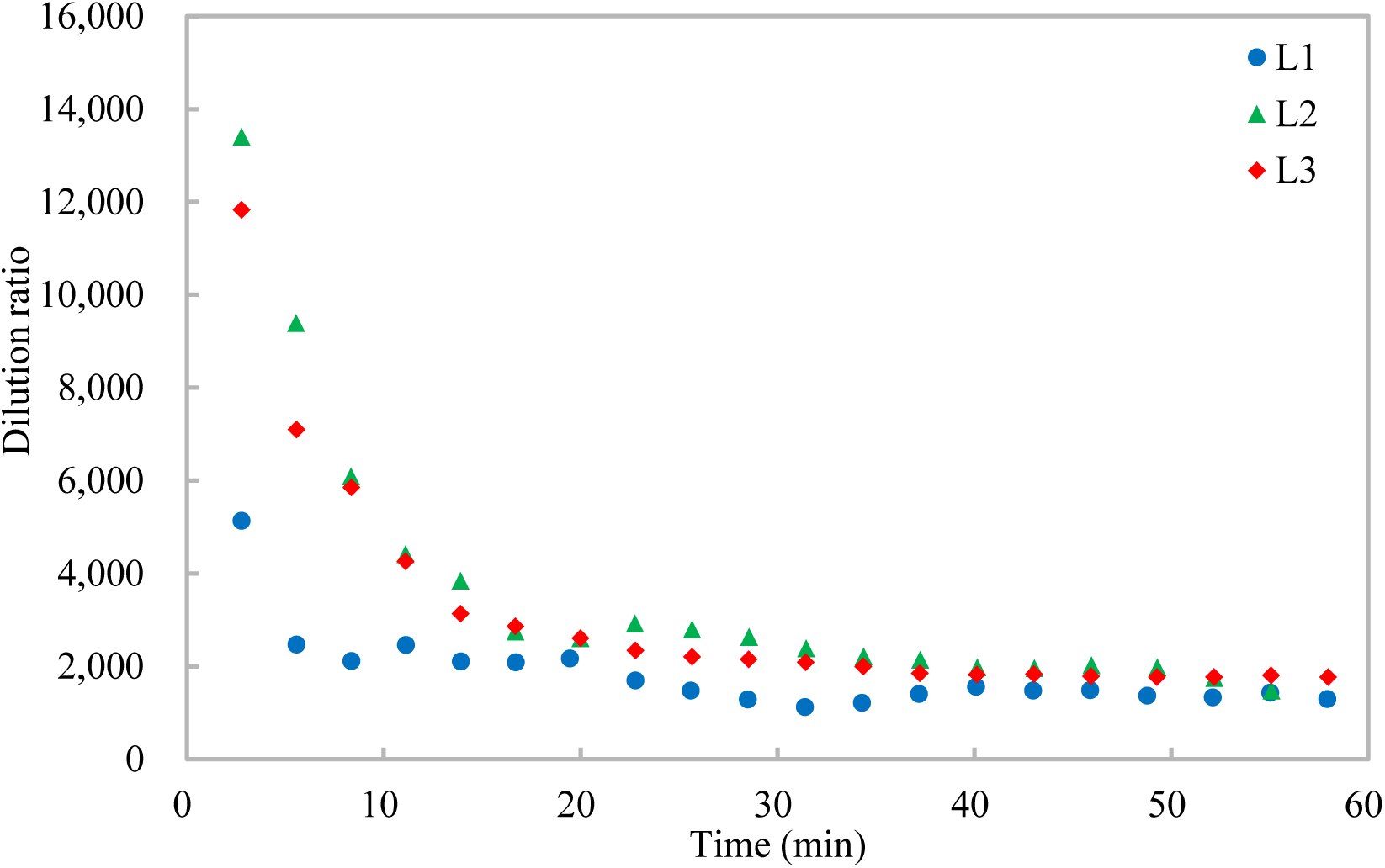
Variations of dilution ratio at different target positions with time.

With the dilution ratio, the airborne infection risks at the three target positions are calculated (Equation 5) (Figures 4 and 5). The quantum generation rate of a COVID-19 infector is assigned to be 142 quanta/h and the breathing rate of a standing susceptible is 0.54 m^3^/h [25]. Two scenarios are considered, one with no masks and the other with surgical masks for both the infector and susceptible. Under Scenario 1 with no masks (*i.e*., with the factor of infectious virus removal of unity) (Figure 4), the airborne infection risks at the three target positions increase with time, and the airborne infection risks at the target positions over the given exposure period (one hour) of L1-L3 are 9.5%, 5.9%, and 6.7% respectively. The large variations of the airborne infection risk indicate that the airborne infection risk should be evaluated both spatially and temporally. Otherwise, the infection airborne risk could be overestimated (*e.g*., for the target position far away from the source, such as L2 and L3) or underestimated (*e.g*., for the target position close to the source, such as L1). When surgical masks with an efficiency of 75% [12] are used by both the infector and susceptible under Scenario 2 (Figure 5), the factor of infectious virus removal is 0.0625 (*i.e*.., (1-0.75)^2^), and the airborne infection risks of all the three target positions are largely reduced to be below 0.7%, indicating that the surgical masks are effective in reducing the cross infections [4]. These results demonstrate the effectiveness of the method proposed in evaluating the airborne infection risk both spatially and temporally.

**Fig.4.**
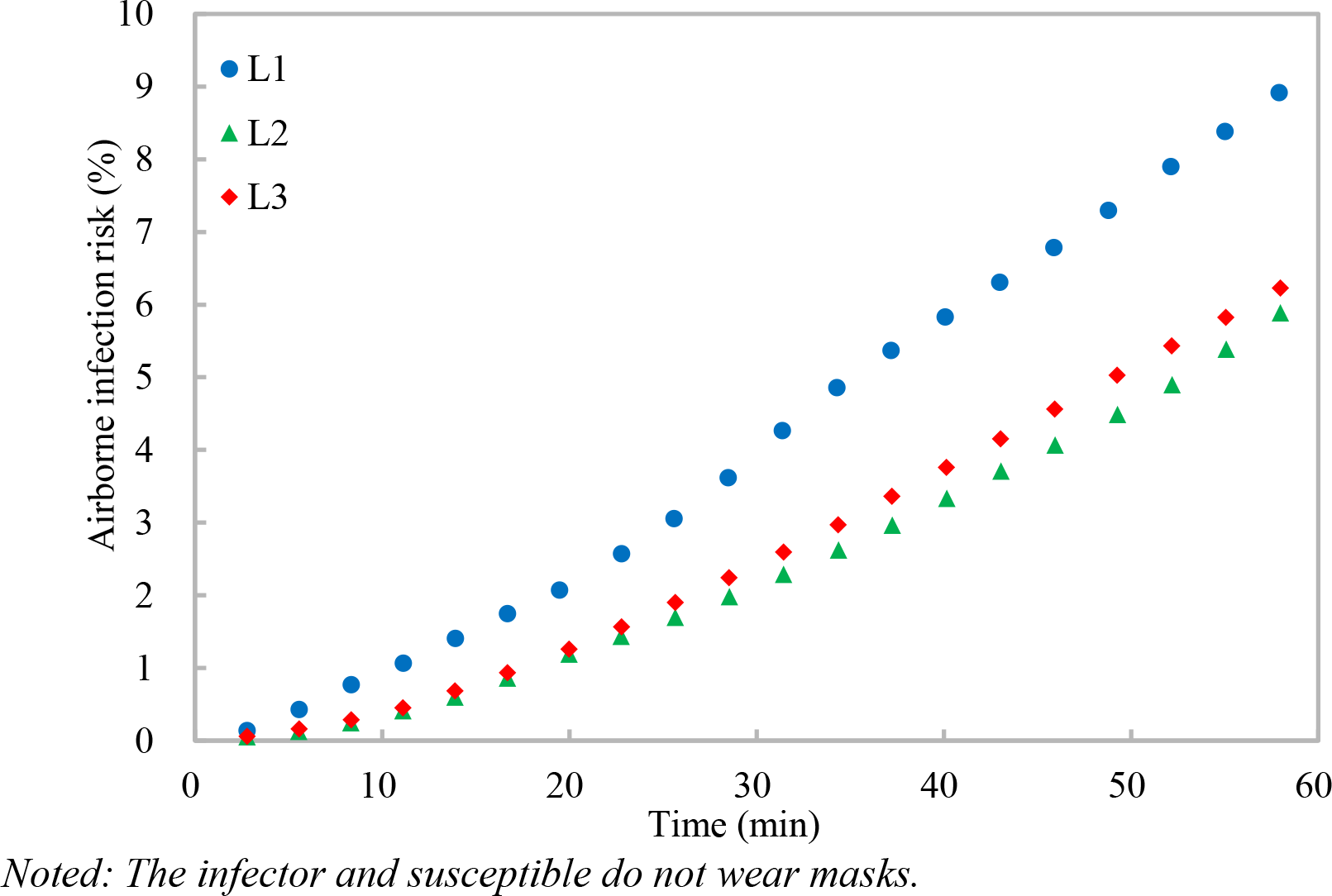
Variations of airborne infection risks at different target positions with time.

**Fig.5.**
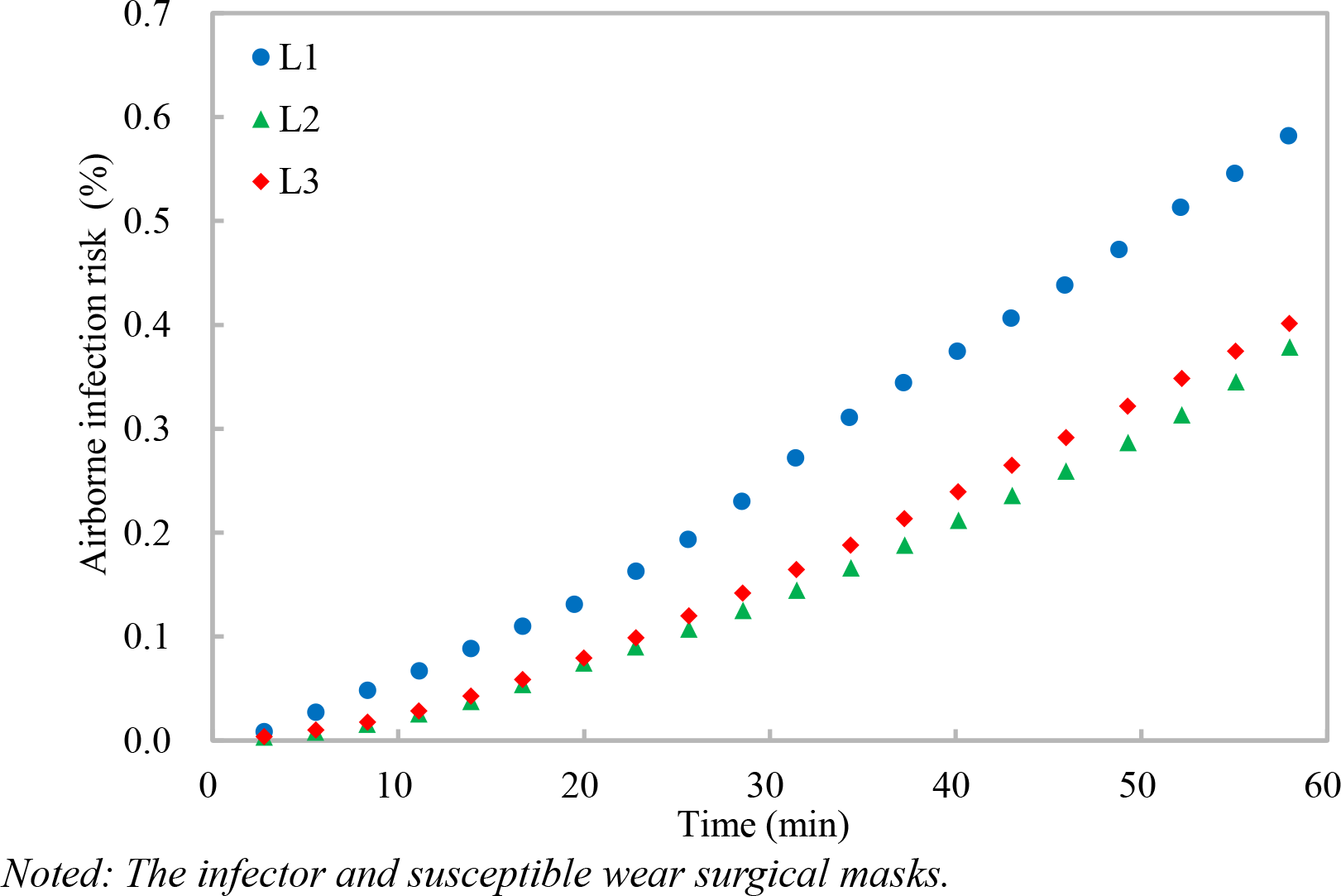
Variations of airborne infection risks at different target positions with time.

## 6. Conclusions

This study proposes an evaluation method for the airborne infection risk based on the concept of dilution. The dilution ratio is used to calculate the inhaled quanta of airborne pathogens with which the airborne infection risk is calculated according to Poisson distribution. The method proposed is benchmarked by the Wells-Riley model under the well-mixed and steady condition, and by a modified Wells-Riley model (*i.e*., rebreathed-fraction model) under the well-mixed and transient condition, which indicates that the method proposed is a thorough expansion of the Wells-Riley model. Compared with the Wells-Riley model, the method proposed has two advantages of 1) evaluation of airborne infection risk for both spatial and temporal resolutions, and 2) convenience in practical applications. Experiments in a mock hospital ward with multiple beds served with displacement ventilation demonstrate that the method proposed effectively evaluates airborne infection risk both spatially and temporally. The method proposed contributes to the reliable evaluation of airborne infection risk and developing effective interventions for reducing cross infections of infectious respiratory diseases (*e.g*., COVID-19).

## Data Availability

All data are available in the manuscript.

## Acknowledgement

The work described in this paper is supported by the National Natural Science Foundation of China (Project No. 51878585). The help of Ms. Yalin Lu in the experiments is highly appreciated.

